# Composite trait Mendelian Randomization reveals distinct metabolic and lifestyle consequences of differences in body shape

**DOI:** 10.1101/2020.09.03.20187567

**Authors:** Jonathan Sulc, Anthony Sonrel, Ninon Mounier, Chiara Auwerx, Eirini Marouli, Liza Darrous, Bogdan Draganski, Tuomas O. Kilpeläinen, Peter Joshi, Ruth J.F. Loos, Zoltán Kutalik

## Abstract

Obesity is a major risk factor for a wide range of cardiometabolic diseases, however the impact of specific aspects of body morphology remains poorly understood. We combined the GWAS summary statistics of fourteen anthropometric traits from UK Biobank through principal component analysis to reveal four major independent axes summarizing 99% of genetically driven variation in body shape and size: overall body size, adiposity, predisposition to abdominal fat deposition, and lean mass. Enrichment analyses suggest that body size and adiposity are affected by genes involved in neuronal signaling, whereas body fat distribution and lean mass are dependent on genes involved in morphogenesis and energy homeostasis. Using Mendelian randomization, we found that although both body size and adiposity contribute to the consequences of BMI, many of their effects are distinct, such as body size increasing the risk of diseases of the veins (*b* ≥ 0.044, p ≤ 8.9*10^-10^) and cardiac arrhythmia (*b* = 0.06, p = 4.2*10^-17^) while adiposity instead increased the risk of ischemic heart disease (*b* = 0.079, p = 8.2*10^-21^). The body mass-neutral component predisposing to abdominal fat deposition, likely reflecting a shift from subcutaneous to visceral fat, exhibited health effects that were weaker but specifically linked to lipotoxicity, such as ischemic heart disease (*b* = 0.067, p = 9.4*10^-14^) and diabetes (*b* = 0.082, p = 5.9*10^-19^). Combining their predicted effects significantly improved the prediction of obesity-related diseases, even when applied out-of-population (p < 10^-10^). The presented decomposition approach sheds light on the biological mechanisms underlying the remarkably heterogeneous nature of body morphology as well as its consequences on health and lifestyle.

## Introduction

Obesity is one of the main risk factors for many non-communicable diseases, such as type 2 diabetes (T2D, reviewed in 1) and cardiovascular diseases (CVD, reviewed in 2). The associated disease risk and underlying biology of obesity are generally studied through the lens of body mass index (BMI, weight [kg] / height^2^ [m^2^]), which uses excess body mass as a surrogate for adiposity. This approximation provides a reasonably accurate predictor at the population level^3, 4^ but is blind to many aspects of body shape and composition that may be critical to disease etiology. Indeed, disease risk and progression have been shown to be affected by the location and type of the adipose tissue in which excess calories are stored^3–6^. Abdominal obesity in particular, usually assessed using waist circumference or waist-to-hip ratio (WHR), is associated with increased disease risk and mortality independent of BMI^3^.

Although the exact mechanisms underlying the consequences of adiposity on health have not been fully elucidated, the predominant hypothesis is that of adipose tissue expandability: that excess calories are preferentially stored in subcutaneous adipose tissue (SAT), which can expand with little to no deleterious impact on health^7, 8^. When its capacity for expansion is exceeded, adipose cells hypertrophy, causing local inflammation, and fat is increasingly stored as ectopic fat in organs and as visceral adipose tissue (VAT) around organs in a process similar to a mild form of lipodystrophy. Ectopic and visceral fat are thought to play a central role in many of the direct consequences of obesity.

Much remains unknown about the genetic and environmental factors affecting body fat distribution and its contribution to health outcomes. Currently available non-invasive imaging techniques such as magnetic resonance imaging (MRI) and dual-energy X-ray absorptiometry (DXA) allow the accurate measurement of adipose mass in different parts of the body and have significantly contributed to our understanding of the impact of different subtypes of adiposity on health^9, 10^. However, such techniques remain costly and are therefore generally restricted to smaller sample sizes.

An alternative approach is the concurrent analysis of multiple traits, leveraging the co-occurring changes in multiple phenotypes to understand the underlying causes and mechanisms. Analysis of variance methods, such as principal component analysis (PCA), have been used to investigate the complex architecture underlying body morphology, revealing the main axes of phenotypic variation and increasing the statistical power to detect novel loci affecting body morphology^11^. While this has improved our understanding of the genetic basis underlying common and distinct components of anthropometric traits, their impact on health and quality of life remains unknown. Other approaches such as clustering and canonical correlation analysis (CCA) have identified single nucleotide polymorphisms (SNPs) associated with healthier metabolic profiles, despite higher BMI and/or body fat percentage^12–15^. However, these hypothesis-driven approaches (i.e. identifying clusters of functionally similar SNPs based on both obesity measures and health outcomes) are not suited to determine the causality of these correlated differences or the directionality of potential causal effects because the SNP groups have different health consequences by construction.

Here we followed a hypothesis-free approach to isolate independent axes of variations in body shape and size and investigated their health consequences. We performed a PCA on GWAS summary statistics of 14 anthropometric traits from the UK Biobank^16^ to extract orthogonal components, each representing different features of body shape (Figure 1). We show that these measures of body shape can be summarized using four principal components (PCs) affecting body size, adiposity, abdominal fat deposition, and lean mass, respectively. Enrichment analyses highlighted differences in the pathways and tissues involved in these composite traits, providing new insight into the underlying biological mechanisms. We then used robust cross-sex Mendelian randomization to assess the impact of these independent components on health and lifestyle. While many health and lifestyle consequences were shared with individual traits, these orthogonal PCs allowed us to better disentangle the independent contributions of different aspects of body shape. The results can be explored using the shiny app which can be downloaded by following the instructions at http://wp.unil.ch/sgg/pca-mr/. Furthermore, the combination of these PCs improved the prediction accuracy of obesity-related diseases.

**Figure 1.**
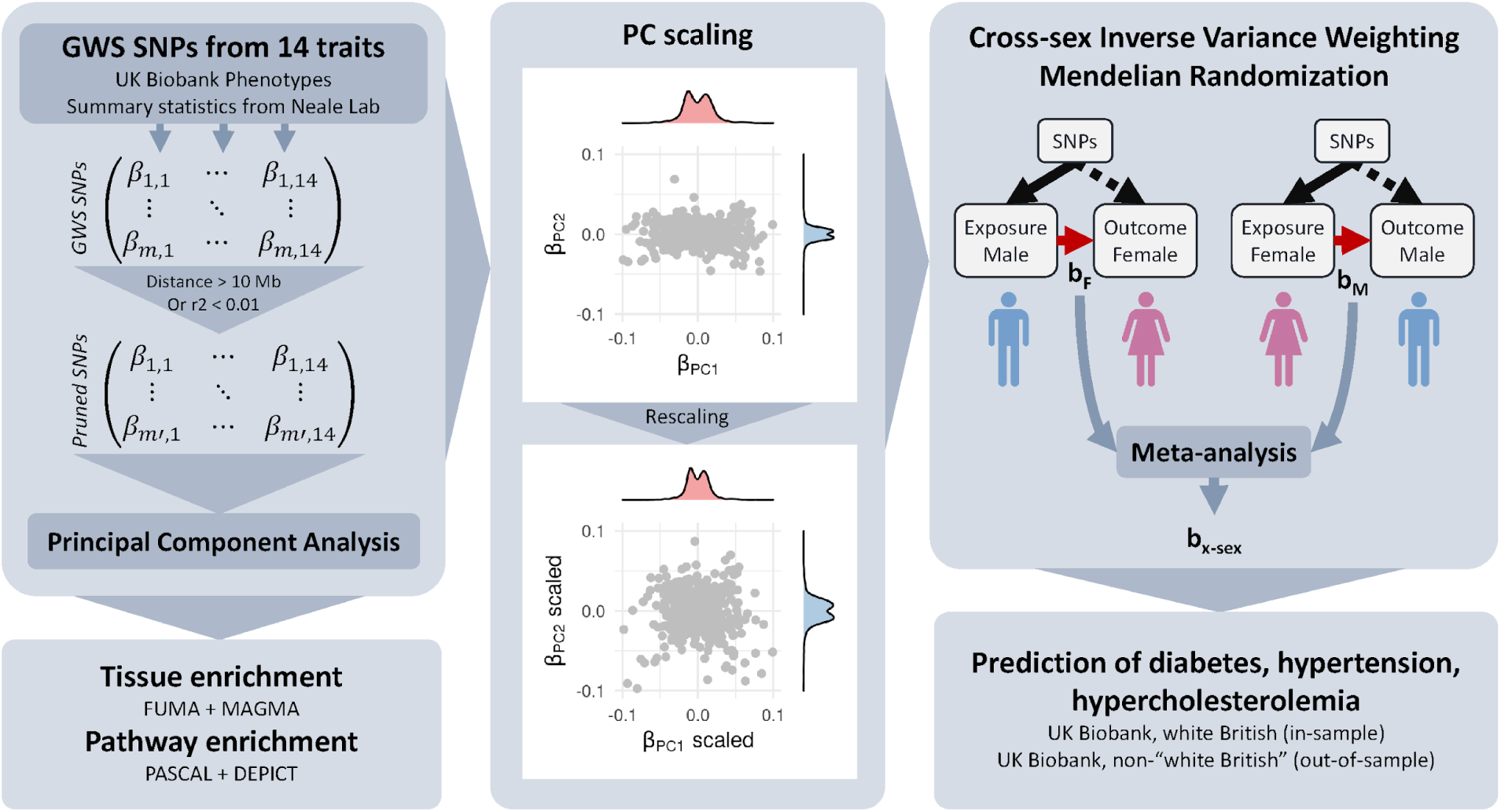
Overview of the methods. Summary statistics for anthropometric traits from the UK Biobank were pruned for independence before being subjected to principal component analysis (PCA). PC-associated SNPs were tested for enrichment in genes expressed in certain tissues or in pathways. The genetic effects on the resulting components were scaled to obtain effect sizes corresponding to a trait with a variance of 1 (standardized). Mendelian randomization was used to determine the impact of these composite traits on lifestyle and health outcomes. Using these effect estimates, individual risk was predicted in the UK Biobank and accuracy compared to BMI and WHR.

## Results

### PCA of genetic effects

A schematic representation of our composite MR analysis framework is shown in Figure 1. We selected 14 anthropometric and bioimpedance-derived traits as the basis for the principal component analysis (PCA), 13 of which were available in the UK Biobank^16^ with genome-wide summary statistics made available by the Neale lab (www.nealelab.is/uk-biobank). Summary statistics for WHR were not available in the UK Biobank, therefore we performed a GWAS in the UK Biobank following the same procedure as that used by the Neale lab. SNPs which were then genome-wide significant (GWS, p < 5*10^-8^) for any trait were pruned to be independent. The resulting SNP x traits matrix of effect estimates was subjected to PCA. The loadings of each principal component (PC) were then rescaled according to the phenotypic correlation between these traits in the UK Biobank so as to obtain standardized effect sizes, i.e. the resulting PC phenotypes would have a variance of 1. SNP-PC associations were then calculated genome-wide and SNPs for each PC were re-pruned individually.

The top four PCs explained more than 99% of the total variance (Figure 2, Supplementary Table 3). PC1 (73.3% variance) represents an overall increase in body size, with positive weights for all traits indicating a slightly disproportionate increase in body mass compared to height, resulting in higher BMI as well. PC2 (19.9%) shows a decrease in height with an increase in fat mass at the expense of lean mass. PC3 (3.8%) is largely BMI- and body fat mass-neutral, decreasing hip circumference, and increasing waist circumference and hence WHR, reflecting a shift in body fat from hips to waist. PC4 (2.4%) resulted in decreased height and increased BMI, with an increase in lean mass at the expense of fat mass.

**Figure 2.**
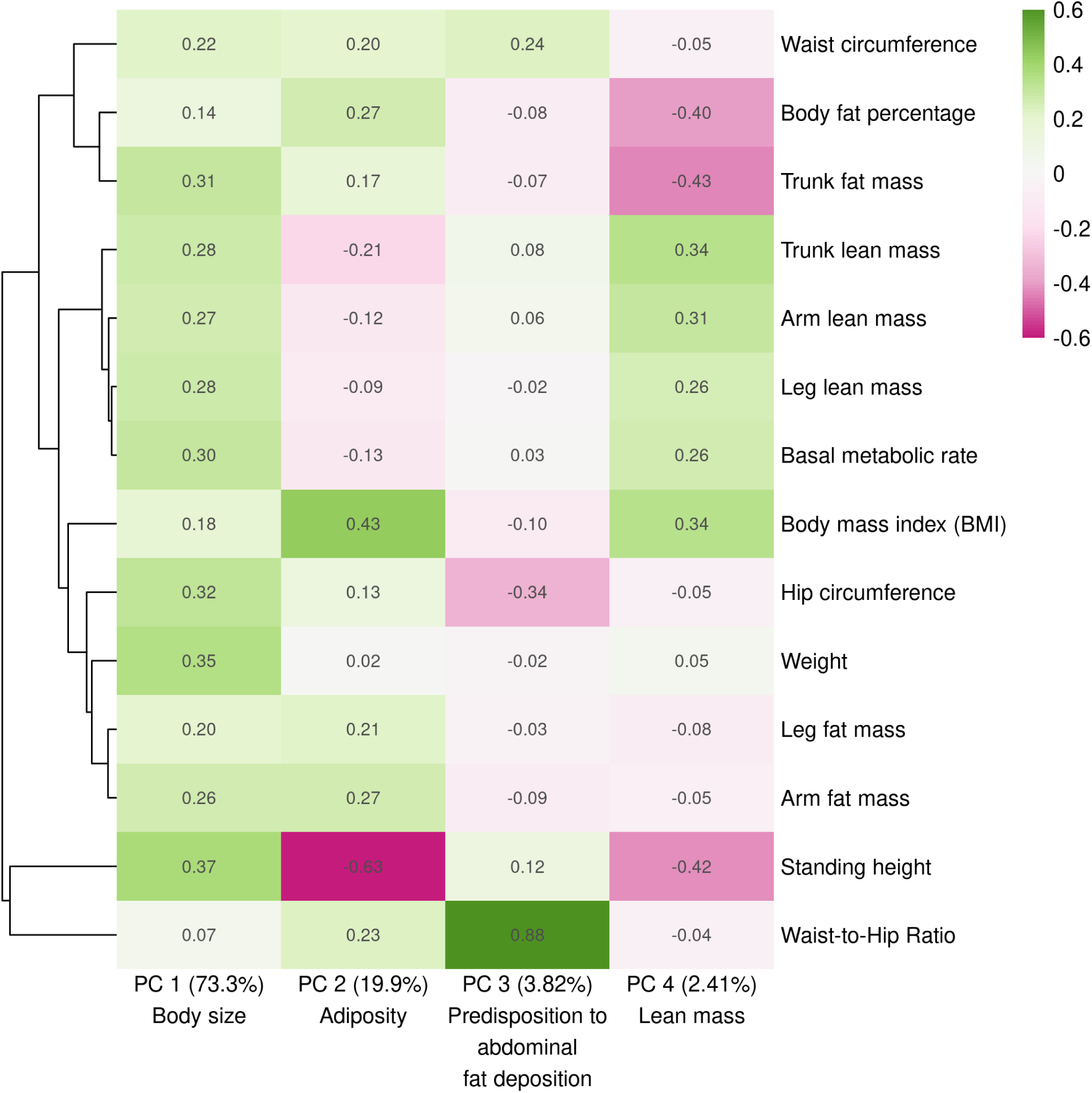
The contributions of each trait to the first four genetic PCs. The explained variance of each PC is included in parentheses along with the descriptive name used in the main text. The loadings presented here are those typically used in PCA, scaled such that the sum of the squared weights is equal to 1 (as opposed to the scaling used to obtain composite traits with a variance of 1). This provides a consistent scale and makes PCs more easily comparable with each other.

Performing this analysis using male- or female-specific summary statistics produced very similar results, though PC3 explained less variance than PC4 in men (Supplementary Tables 4-5, Supplementary Figure 1). PCs were also robust to changes in the selection of specific traits. For example, excluding WHR yielded effect estimates which were highly correlated with those obtained using the full set of traits (all |r| > 0.99, see Supplementary Figure 2), though the altered explained variance also resulted in the reordering of PCs 3 and 4.

PCs 1-3 were highly correlated with weight, BMI, and WHR, respectively, both phenotypically (r ≥ 0.70, Supplementary Figure 14) and in terms of their causal effects on the tested outcomes. The (genetically) orthogonal nature of PCs nevertheless resulted in much lower phenotypic correlation with each other than between traits (e.g. the phenotypic correlation between PC1 and PC2 was only 0.36 while that between weight and BMI was 0.90, Supplementary Figure 15, Supplementary Table 11). Note that the phenotypic realization of PCs are not orthogonal, and their correlation is therefore not zero, due to differences between their environmental and genetic correlation. However, the MR-derived causal effect estimates of PCs are independent and therefore additive (though they may still be correlated), which is not the case for individual traits.

For brevity and clarity, we mainly describe the PCs in comparison with BMI and occasionally weight or WHR. Other traits had causal effects similar to or weaker than these traits (e.g. the effects of body fat percentage and BMI on disease had a correlation coefficient r = 0.95, p = 7.6*10^-43^) or were less relevant to obesity-related health outcomes (e.g. height).

We found 615 independent GWS SNPs associated with PC1 (body size), 641 with PC2 (adiposity), 354 with PC3 (predisposition to abdominal fat deposition), and 610 with PC4 (lean mass). Among these, respectively 3, 83, 137, and 330 SNPs did not reach genome-wide significance for any individual trait. For comparison, BMI had 532 independent GWS SNPs.

### Tissue/pathway enrichment

We tested the PCs for enrichment of genes expressed in specific tissues using the MAGMA method^17^ through the FUMA interface^18^ in both GTEx v8^19^ and brain development and aging data from BrainSpan^20^. We also tested for their enrichment of molecular pathway terms from the DEPICT dataset^21^ using PASCAL^22^. The tissue-enrichment results of the four PCs as well as weight, BMI and WHR are shown in Figure 3 and listed in Supplementary Tables 6–9.

**Figure 3.**
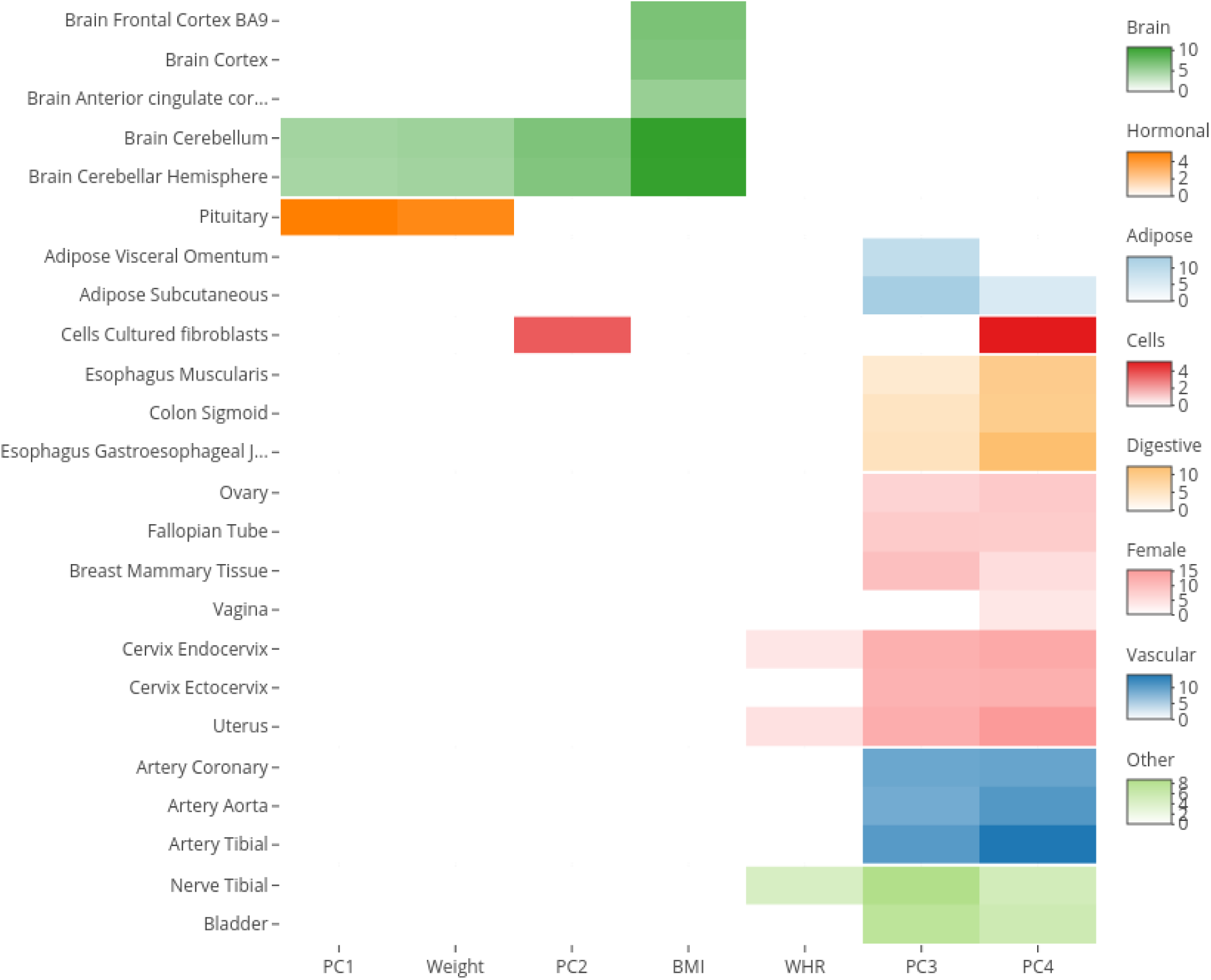
Body size and accumulation of body fat were mainly enriched for genes expressed in the brain, while the others were enriched for a broader range of tissues. Enrichment of traits and principal components (PCs) for tissue-specific gene expression (negative log 10 p-values). Genome-wide SNP effect p-values were analyzed using MAGMA on GTEx v8 data (54 tissues). Results not significant after Bonferroni correction are masked in white. Traits with no significant enrichment results are hidden for clarity (full results are available in Supplementary Table 6).

Loci associated with both PC1 (body size) and PC2 (adiposity) were mainly enriched for genes expressed in the cerebellum (p ≤ 2.2*10^-5^) and the pituitary gland (p ≤ 5.7*10^-4^). Using data from BrainSpan^20^, we found PC2 to be further enriched for genes expressed in the brain specifically during the early to late mid-prenatal phases of development (p ≤ 6.1*10^-4^).

PC3- (predisposition to abdominal fat deposition) associated SNPs were harboured by genes most enriched for expression in SAT (p = 3.0*10^-14^), followed by VAT (p = 4.2*10^-10^), female reproductive tissues (breast mammary tissue, ecto- and endocervix, and uterus, all p ≤ 2.4*10^-10^), nerves (tibial, p = 2*10^-15^), arteries (p ≤ 2.2*10^-9^), and digestive system (p ≤ 8.8*10^-5^). Note that using sex-specific (e.g. male-specific) summary statistics and PC loadings with the same gene expression datasets produced a similar enrichment for female-specific tissues. Data from BrainSpan showed PC3 to be enriched for genes expressed in the late prenatal brain (p = 9.3*10^-8^).

PC4 (lean mass) showed similar enrichment to that of PC3, though stronger for the digestive system (p ≤ 1.6*10^-10^) and some female reproductive tissues (uterus, ecto- and endocervix, all p ≤ 1.3*10^-12^),and weaker for adipose tissue and tibial nerve. PC4 was also enriched for genes expressed in the prostate (p = 2.9*10^-4^). In BrainSpan, PC4 also showed enrichment for genes expressed prenatally in the brain.

Loci associated with BMI showed slightly stronger enrichment than PCs 1 and 2 for genes expressed in the cerebellum, as well as other areas of the adult brain, including the basal ganglia, hippocampus, hypothalamus, amygdala, and frontal cortex. They were also enriched for genes expressed in the mid-prenatal brain, similar to that found for PC2, if slightly weaker.

The analysis of molecular pathways (Supplementary Table 10) showed qualitatively similar enrichment for PCs 1 and 2, weight and BMI. These were mostly terms related to the brain, synapses, behaviour, or learning. PC3 had the most numerous enriched terms and the strongest enrichment overall. Most of the enriched terms were not related to brain function, but to embryogenesis and morphology, with many others specifically concerning adiposity, metabolism, and glucose homeostasis. Other terms were related to vascular or heart function, or hormones. PC4 showed some overlap in terms with PC3, mainly in terms related to embryogenesis and morphology.

### Mendelian Randomization analysis

#### Cross-sex MR to avoid bias from sample-overlap

We used inverse-variance weighted (IVW) Mendelian randomization (MR) to test for causal effects of the 14 anthropometric traits and PCs on disease outcomes, continuous measures of health, lifestyle factors, and diet, as well as the reverse. To avoid bias in the MR estimates due to sample overlap, we used sex-specific effects from opposite sexes for exposures and outcomes (male exposure - female outcome and female exposure - male outcome, using the same combined-sex PC loadings for exposure PCs). This produces unbiased estimates of the sex-specific causal effects in the sex used in the outcome, provided the strength of the instruments’ association with the exposure was not different between sexes. The two cross-sex causal effect estimates (female-to-male and male-to-female) were then meta-analysed using inverse-variance weighting. Where the strength of the instruments’ association with the exposure differed between sexes, namely for WHR and PC3 (abdominal fat distribution), the meta-analyzed causal effect estimates may be biased (generally towards the null). Although the effect size estimates may be slightly over- or underestimated, this should not affect the type I error rate (see Methods).

#### Disease risk

PC1 (body size) increased the risk of many diseases (Figure 4). An increase of one standard deviation (SD) increased the absolute risk of diabetes by 1.7% (95% CI: 1.3–2.1), that of hypertension by 2.3% (95% CI: 1.4–3.2), as well as many other diseases, such as nerve disorders, diseases of the veins and circulatory system, and prolapsed disc (Supplementary Table 12). Although it also increased the risk of cardiac arrhythmias by 0.93% (95% CI: 0.71– 1.1), it did not significantly affect the risk of hypercholesterolemia or heart disease.

**Figure 4.**
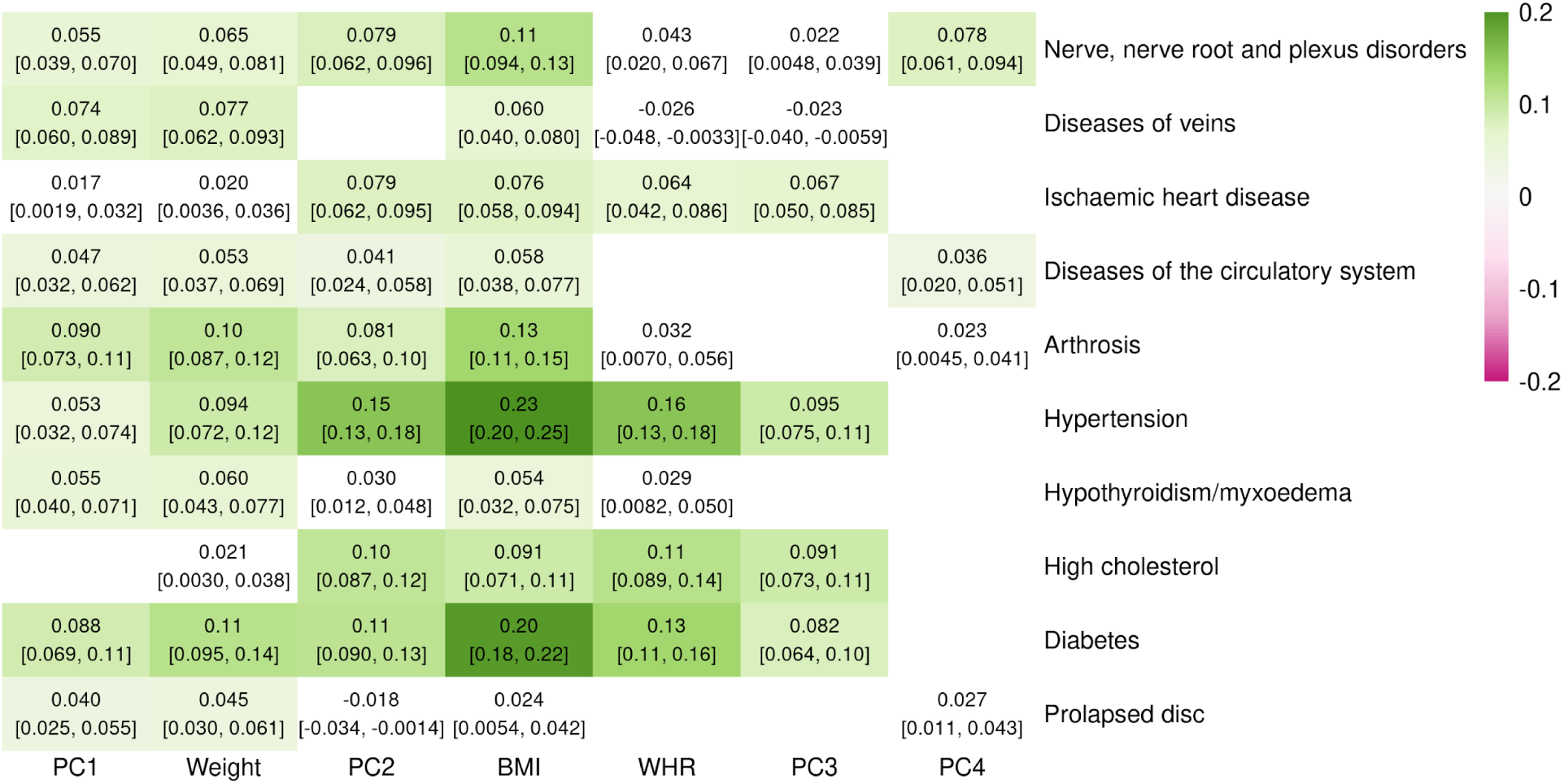
Single and composite traits increase the risk of multiple diseases. MR causal effects of traits and principal components (PCs) on a selection of diseases on a standardized scale. The 95% confidence interval of the effect is indicated in brackets. Effects which were not significant at the Bonferroni-corrected threshold (p < 4.3*10^-5^) are colored in white. The full list of effects can be found in Supplementary table 12.

PC2 (adiposity) had much stronger effects on many obesity-related diseases (Figure 4), where a 1 SD increase also increased the absolute risk of diabetes by 2.1% (95% CI: 1.7–2.5), hypertension by 6.8% (95% CI: 5.8–7.7), as well as hypercholesterolemia by 3.4% (95% CI: 2.8–4.0) and ischaemic heart disease (IHD) by 1.8% (95% CI: 1.5–2.2). The risk of many other diseases, such as arthrosis and diseases of the nervous system were also increased (Supplementary Table 12).

PC3 (predisposition to abdominal fat deposition), despite being weight- and BMI-neutral, was a risk factor for many of the same obesity-related diseases as PC2 (Figure 4, Supplementary Table 12). A 1 SD increase in PC3 increased the absolute risk of diabetes by 1.6% (95% CI: 1.2–1.9), hypertension by 4.2% (95% CI: 3.3–5.0), hypercholesterolemia by 3.0% (95% CI: 2.4–3.6), and IHD by 1.6% (95% CI: 1.2–2.0).

PC4 (lean mass) had few consequences on health, only significantly increasing the risk of nerve disorders and diseases, carpal tunnel syndrome, and joint disorders (Figure 4, Supplementary Table 12).

For comparison, a 1 SD increase in BMI (5.1 kg/m^2^ in women, 4.2 kg/m^2^ in men) increased the risk of diabetes by 3.8% (95% CI: 3.4–4.2), hypertension by 9.9% (95% CI: 8.8–11.0), hypercholesterolemia by 3.0% (95% CI: 2.3–3.7), and IHD by 1.8% (95% CI: 1.4–2.2). Weight and WHR had similar, if somewhat weaker, effects (see Supplementary Table 12). Although in many cases these effects exceed those of individual PCs, they remain less than the cumulative (summed) effects of the four PCs.

#### Continuous health outcomes

Many continuous health indicators were affected by both PCs and traits, in a manner largely consistent with expectations (Supplementary Table 13).

Consistent with the increased risk of diabetes, PCs 1-3 all increased the levels of glycated hemoglobin in blood, though the effect was strongest for PC2 (b = 0.13, 95% CI: 0.11–0.15). Glucose levels were similarly affected, though the effects were weaker. All three PCs also increased triglyceride levels (*b* between 0.095 and 0.24) and decreased HDL cholesterol (b between −0.22 and −0.16), but only PC1 decreased LDL cholesterol (b = −0.076, 95% CI: - 0.094– −0.059) and total cholesterol. They all increased blood pressure (systolic and/or diastolic), with the strongest effects from PC2 (b > 0.15). Plasma concentrations of several liver enzymes, such as *γ*-glutamyltransferase (GGT) and alanine aminotransferase (ALT), were found to be increased as well, possibly indicating liver damage.

PCs 1 and 2 also increased levels of cystatin C and decreased albumin, possibly indicative of impaired kidney function^23^, whereas the effects of PC3 were in the opposite direction (decreasing cystatin C and increasing albumin). C-reactive protein (CRP) was also increased by PCs 1 and 2 (b_1_ = 0.17, b_2_ = 0.25) but unaffected by PC3. PCs 1-3 all strongly decreased levels of sex hormone-binding globulin (SHBG) with the strongest effects from PC2 (b = −0.19, 95% CI: −0.21– −0.16), as well as testosterone where the effects of PCs 1 and 3 were stronger (b_1_ = - 0.065, b_3_ = −0.058).

PC4 mainly increased creatinine levels while decreasing CRP (b = −0.090, 95% CI: −0.11– - 0.067) and the maximum heart rate during fitness test. The levels of triglycerides and the liver function markers GGT and ALT were slightly decreased but only alkaline phosphatase reached Bonferroni-corrected significance (b = −0.068, 95% CI: −0.09– −0.046).

The effects of BMI were comparable to a combination of PCs 1 and 2, increasing levels of glycated hemoglobin (b = 0.2) and glucose (b = 0.12), as well as increasing the levels of triglycerides while decreasing both HDL (b = −0.29) and LDL (b = −0.059) cholesterol. Systolic and diastolic blood pressure was also increased, as were levels of CRP (b = 0.33) and liver function markers such as ALT and GGT. SHBG (b = −0.24) and testosterone (b = −0.11) were also decreased.

#### Lifestyle factors

PC1 slightly reduced SES, as shown by increased TDI (b = 0.033, 95% CI: 0.017–0.049). This was accompanied by a longer working week, as well as an increase in smoking and alcohol consumption, particularly spirits (Figure 5, Supplementary Table 14). The duration and frequency of physical activity, as well as walking pace, were all decreased in favor of increased time spent using the computer. PC1 also increased daytime dozing and napping but decreased snoring.

**Figure 5.**
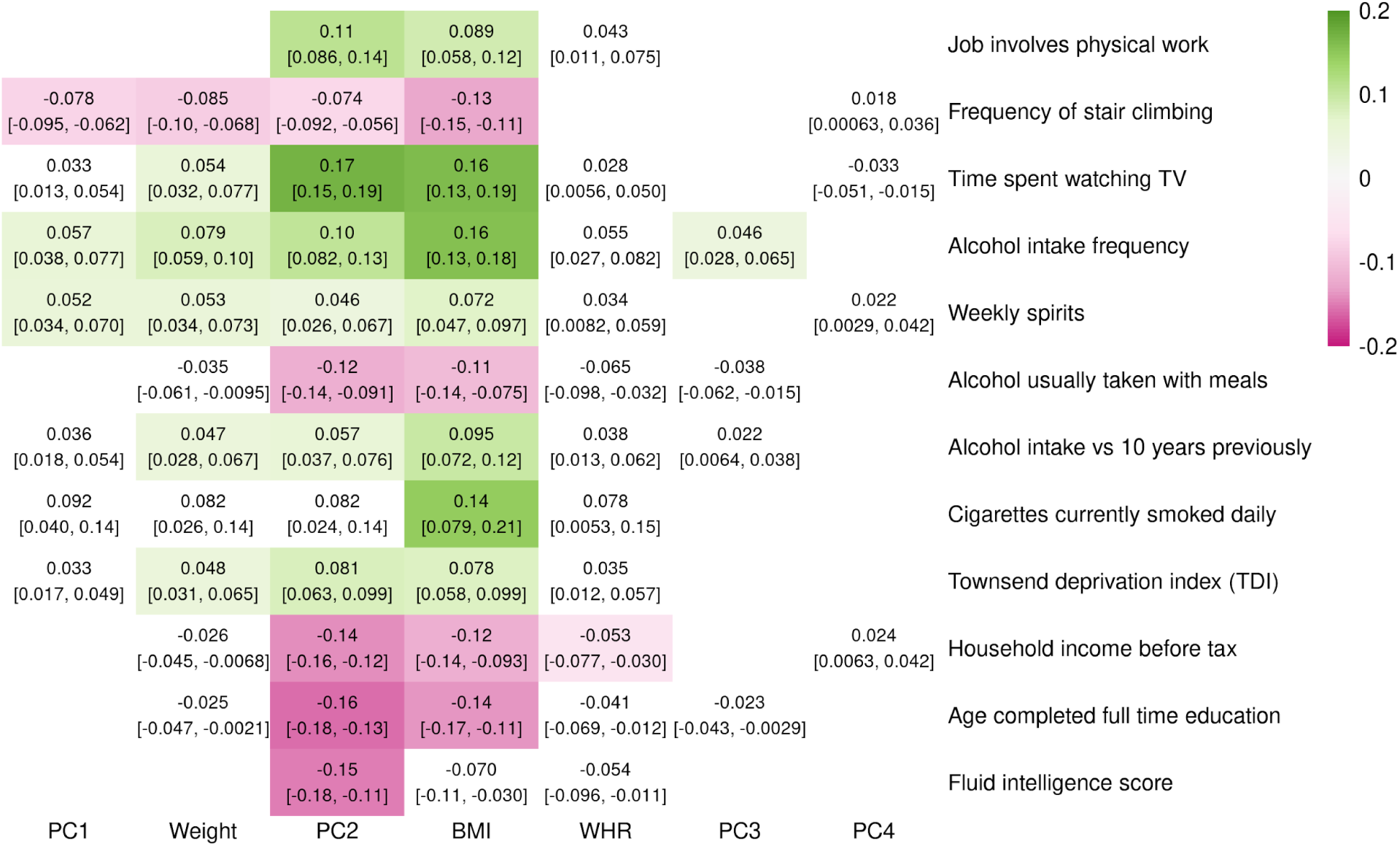
Single and composite traits affect many aspects of lifestyle. MR causal effects of traits and principal components (PCs) on a selection of lifestyle factors on a standardized scale. The 95% confidence interval of the effect is indicated in brackets. Effects which were not significant at the Bonferroni-corrected threshold (p < 7.1*10^-5^) are colored in white. The full list of effects can be found in Supplementary table 14.

The effects of PC2 on SES were similar but much more pronounced (Figure 5, Supplementary Table 14), not only associated with increased TDI (b = 0.057, 95%CI: 0.037– 0.076) and the likelihood of having a job involving heavy physical work (b = 0.11, 95% CI: 0.086–0.14), but strongly linked to decreased income, fluid intelligence score, and education (all b < −0.14). Accompanying these were lifestyle changes similar to those of PC1, increasing smoking and the frequency of alcohol consumption, with a decrease in wine in favor of spirits and alcohol being taken more often taken outside of meals. Although PC2 increased the duration of walks (b = 0.05, 95% CI: 0.031–0.069) and vigorous activity, the duration of walking for pleasure was decreased (b = −0.044, 95% CI: −0.062– −0.025), as were several other measures of physical activity, namely the frequency of stair climbing and the usual walking pace. Unlike PC1, PC2 decreased the time spent using the computer in favor of time spent watching TV.

PC3 increased alcohol intake frequency (b = 0.046, 95% CI: 0.028– 0.065) and napping during the day (b = 0.035, 95% CI: 0.018–0.053) (Figure 5, Supplementary Table 14).

PC4 only associated with an increased length of the working week (b = 0.043, 95% CI: 0.024–0.062) (Supplementary Table 14).

The effects of BMI on lifestyle were most similar to those of PC2, linked to decreased SES (b_TDI_ = 0.078) and physical activity while increasing smoking and alcohol consumption.

#### Diet

PC1 was associated with greater reported variation in diet (b = 0.052, 95% CI: 0.034–0.069) and increased consumption of healthy foods such as fresh fruit, vegetables, and water (Supplementary Table 15). Consumption of coffee was also increased (b = 0.087, 95% CI: 0.068–0.11).

PC2 was similarly associated with reportedly increased variation in diet (b = 0.084, 95% CI: 0.065–0.10), as well as salt added to food (b = 0.045, 95% CI: 0.026–0.064). Other changes in diet reflect increased consumption of cheaper meats, namely pork and poultry, while decreasing intake of grain products (bread and cereal), cheese, and dried fruits (Supplementary Table 15).

PC3 was associated with increased bread consumption (b = 0.035, 95% CI: 0.017–0.053).

PC4 increased fruit intake, both fresh and dried (b > 0.041), while decreasing processed meat intake (Supplementary Table 15).

Effects of BMI on diet were similar to those of PC2, such as increased variation in diet (b = 0.12, 95% CI: 0.097–0.14) and decreased intake of grain products and cheese (Supplementary Table 15). In addition, some effects were similar to those of PC1, including increased vegetable and fruit consumption, as well as coffee intake (b = 0.11, 95% CI: 0.080–0.13).

#### Sex-specific effects

The genetic effects of the selected IVs on PCs 1, 2, and 4, as well as BMI and weight, were not significantly different between men and women, which made it possible to obtain unbiased sex-specific causal effects (see Methods). Those of PC3 (and WHR) were stronger in women (p_PC3_ = 6.9*10^-46^, p_WHR_ = 1.4*10^-159^) and were unsuitable for this purpose. Full results are available in Supplementary Tables 16–23 and illustrated in Supplementary Figures 100–121.

None of the PCs had significantly different effects on diet and lifestyle in men and women, though PC2 did have a tendency for stronger effects on disease risk in men (TLS slope = 1.27), mainly driven by diabetes (b_m_ = 0.15, b_f_ = 0.080, p_diff_ = 1.8*10^-3^) and IHD (b_m_ = 0.11, b_f_ = 0.059, p_diff_ = 4.7*10^-3^). However, many blood molecular traits and other continuous health outcomes were differently affected by PCs in men and women.

Testosterone in particular was differently affected between sexes, strongly decreased in men by both PC1 (b = −0.15) and PC2 (b = −0.18), whereas in women it was unaffected by PC1 and increased by PC2 (b = 0.082). SHBG was similarly affected by PC2 in both sexes, but the decrease caused by PC1 was twice as strong in women (b_m_ = −0.084, b_f_ = −0.17, p_diff_ = 1.5*10^-4^).

Other differences include PC1 causing a stronger decrease in albumin in women (b_m_ = - 0.095, b_f_ = −0.17, p_diff_ = 6.0*10^-5^) and increasing glycated hemoglobin only in men (b = 0.12, p_diff_ = 5.8*10^-5^). The effects of PC2 in men were stronger for ALT, GGT, and pulse rate, and an increase in total protein was only seen in men.

The effects of BMI were again similar to those of PCs 1 and 2. The increase in glycated hemoglobin was also stronger in men and was accompanied a larger increase in glucose as well (b_m_ = 0.16, b_f_ = 0.083, p_diff_ = 5.2*10^-4^). LDL was decreased only in men (b = −0.099). The opposite effects on testosterone levels were also observed for BMI (b_m_ = −0.24, b_f_ = 0.072).

#### Bi-directional MR

In addition to determining which environmental factors may affect body shape, we were interested in feedback loops. The stringency of the selection criteria for IVs, namely that they be GWS for the sex used as exposure, resulted in few exposures significantly affecting the body traits or PCs. Several exposures had unrealistically large estimated effect sizes (> 0.4), likely due to confounding, and are not considered here. For example, forced vital capacity, which is dependent on lung—and therefore body—size, showed strong bi-directional associations with most body traits. The main results are summarized here, full results are listed in Supplementary Tables 24–27.

While PC1 reduced cholesterol, it was in turn reduced by LDL and total cholesterol (b ≤ - 0.068), as well as apolipoprotein A and diagnosis of hypercholesterolemia. PC1 also showed a positive feedback loop with cystatin C but a negative one with IGF-1, reducing IGF-1 levels in blood (b = −0.13, 95% CI: −0.16– −0.11) but being increased by it (b = 0.042, 95% CI: 0.024–0.061).

PC2 showed a positive feedback loop with CRP and mutual negative loop with creatinine. PC2 was also reduced by IGF-1 (b = −0.071, 95% CI: −0.089– −0.052).

PC3 was decreased by hypertension (b = −0.07, 95% CI: −0.11– −0.035), forming a negative feedback loop. It also formed a positive feedback loop with diabetes (as did WHR), although this seems to be due to a negative effect of diabetes on hip circumference (HC, b = −0.16, 95% CI: - 0.26– −0.067) rather than an increase in waist circumference (WC). PC3 also had a positive feedback loop with triglycerides (from decreased HC), and mutual negative effects on apolipoprotein A and HDL cholesterol (from decreased WC). We also found the negative effects of PC3 on testosterone and SHBG to be reciprocal (b ≤ −0.058).

PC4 had a positive feedback loop with creatinine and was also increased by IGF-1 (b = 0.12, 95% CI: 0.11–0.14). SHBG and testosterone both decreased PC4, though for testosterone this appears to have been driven by an effect in women only (b = −0.09, 95% CI: −0.11– −0.065).

BMI increased CRP in a positive feedback loop, similar to that of PC2, and reciprocal negative effects with LDL and total cholesterol (b ≤ −0.035), similar to PC1.

#### DXA-based measures

DXA and MRI technologies provide increased accuracy in measurements of body composition and have been suggested to provide a clearer picture of obesity and its consequences on health^24^. We were able to impute many such traits based on the subset of ∼5,000 UK Biobank participants with these phenotypes (Supplementary Table 2). Several of these, including both trunk and android tissue fat percentages (both predicted with r^2^ above 0.75), had effects on disease comparable to BMI and PC2 (correlation r ≥ 0.97) but with a tendency for slightly larger effects (TLS slopes of 1.17 and 1.16, respectively). However, with fewer GWS SNPs (∼28% fewer than BMI), these phenotypes revealed no additional diseases affected by obesity (see Supplementary Table 28).

#### Prediction of disease risk

In addition to disentangling the causal relationship between body shape and health, these PCs can be used to predict disease outcomes. Furthermore, given the independence of the estimated PC effects, these can be used in a weighted linear combination to improve prediction of specific diseases, based on the estimated causal effects. We compared the accuracy of the PC-based estimator with both BMI and WHR for the most common obesity-related diagnoses in our dataset, namely diabetes, high cholesterol, and hypertension, in the same sample using receiver operating characteristic (ROC) curves.

As expected, both BMI and WHR were effective in predicting these obesity-related diseases, as indicated by areas under the ROC curve (AUC) ranging from 0.62 to 0.74. BMI was more accurate in predicting hypertension (0.66 vs. 0.64, p = 1.3*10^-72^), while WHR was more accurate for high cholesterol (0.65 vs. 0.62, p = 4.9*10^-101^) and slightly more for diabetes (0.74 vs. 0.73, p = 3.0*10^-5^). In all cases, the PC-based predictors significantly outperformed both BMI and WHR with AUCs of 0.68 for hypertension, 0.67 for high cholesterol, and 0.78 for diabetes (all p < 2.3*10^-183^, Supplementary Figure 129–130, Supplementary Table 29).

To explore the transferability of PCs, we replicated these results out-of-sample in 76,756 non-”white British” UK Biobank participants, who had therefore been excluded from the original summary statistics. Despite the considerable potential for disparities between these samples, the AUCs for high cholesterol and hypertension were almost exactly identical to those obtained above (Figure 6, Supplementary Figure 131, Supplementary Table 29), with PC-based predictors significantly outperforming either trait (p ≤ 2.0*10^-43^). The accuracy of PCs and BMI in predicting diabetes out-of-sample decreased to 0.75 and 0.68, respectively, and although WHR was no less accurate out-of-sample (AUC = 0.74) it remained less accurate than PCs (p = 5.1*10^-11^).

**Figure 6.**
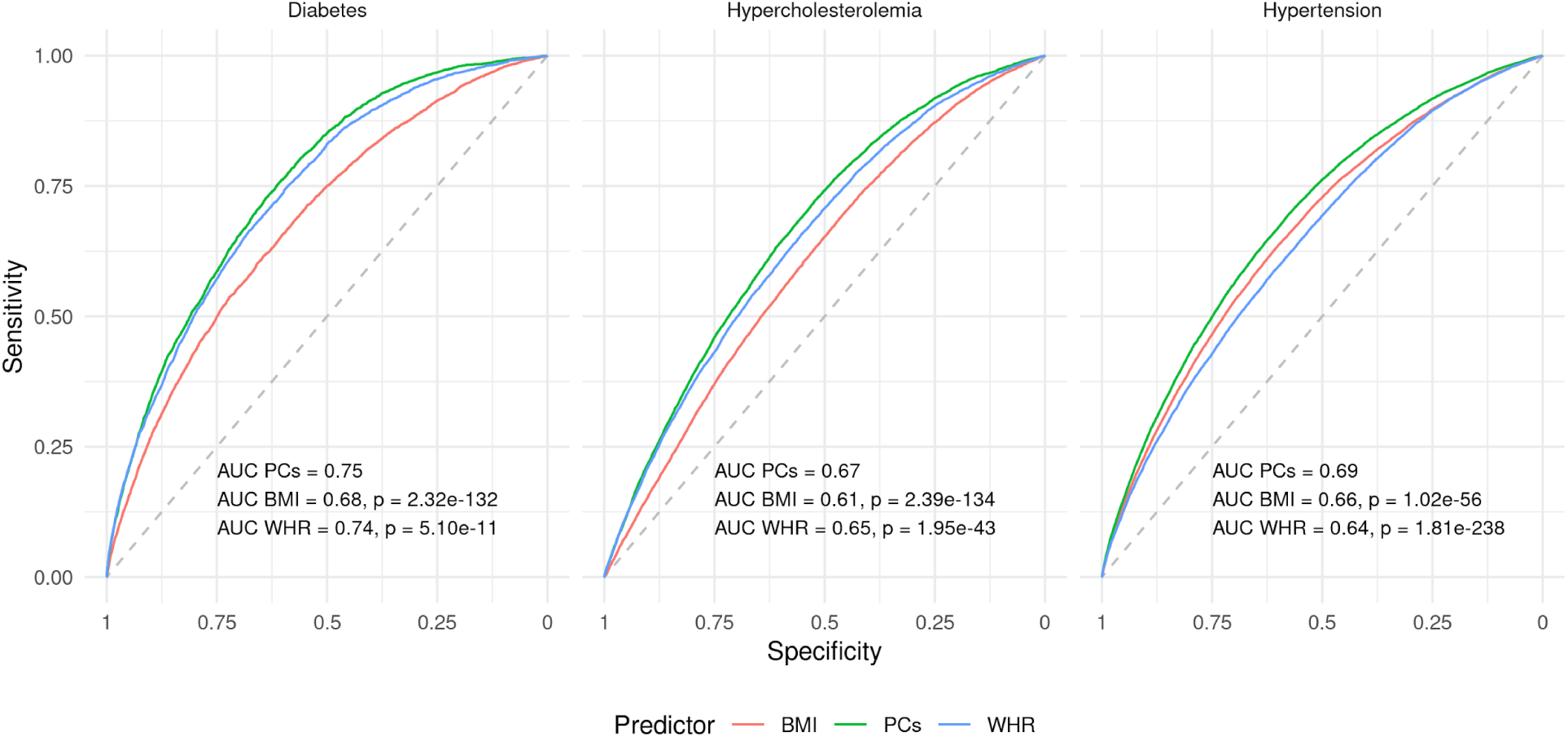
PCs improve prediction of obesity-related diseases out-of-population. ROC curves for PC-, BMI-, and WHR-based prediction of diabetes, hypercholesterolemia, and hypertension out-of-sample/-population. The indicated p-values for the difference between the PC- and single trait-based curves were obtained using the DeLong method.

## Discussion

By combining multiple traits through PCA, we found that more than 99% of genetically-determined variation in body shape and size (as defined by the 14 selected traits) can be summarized using four PCs affecting (1) body size, (2) adiposity, (3) predisposition to abdominal fat deposition, and (4) lean mass. PCs 1–3 showed some similarity with weight, BMI, and WHR, respectively, and the latter two especially have been widely studied in the context of obesity. However, these traits are highly correlated and partially redundant, making it difficult to understand their individual contributions to health outcomes, whereas PCs are orthogonal by design and their effects additive. The PCs obtained here also share some similarity with the average PCs (avPCs) obtained by Ried et al.^11^ by meta-analyzing PCs from individual-level phenotypic data. Specifically, avPC1 increased all body measures (though the impact of height was weaker than for PC1); avPC2 showed opposing directions for height and BMI/WHR (though WHR had much more importance than BMI); avPC3 was dominated by WHR (however both height and BMI were significant contributors as well); and loadings for avPC4 showed an increase in weight and BMI despite decreased waist and hip circumference, suggesting increased density of body mass and consistent with decreased body fat percentage. The robustness of the obtained PCs to changes in the trait selection or the use of male- or female-specific data supports the hypothesis that these represent true biological mechanisms underlying the shared variance across these anthropometric traits. Furthermore, the molecular basis of PCs 3 and 4 in particular appear to be more homogenous, as shown by enrichment for genes expressed in many tissues, such as SAT and VAT but also in the digestive, reproductive, and vascular systems, which were not picked up by individual traits. This is also reflected in the much broader spectrum of pathways enriched only for these PCs.

Leveraging the orthogonality of PCs, we can dissect the etiology of obesity-related health and lifestyle consequences by comparing the effects of different PCs. Both PCs 1 and 2 embody the excessive accumulation of body mass and their enrichment for genes expressed in the brain and involved in neuronal signaling is consistent with findings from BMI^25^ suggesting that behavioral changes are likely one of the major factors underpinning heritable susceptibility to obesity (reviewed in 26). Shared effects between PC1 and PC2 highlight diseases whose etiology involves elements common to both, such as metabolic overexertion in the case of diabetes^27^ or the physical burden of a larger body in the case of arthrosis^28^. Differences between their effects can provide insight into disease etiology which single traits cannot. For example, Hyppönen et al.^29^ performed a phenome-wide MR analysis to assess the effects of BMI on a number of diseases and found that BMI increased the risk of IHD, cardiac ar-/dysrhythmia, and diseases of the veins (phlebitis and thrombophlebitis), among others. However, our results show that these do not all arise through the same mechanisms, IHD being increased by PCs 2 and 3 but not PC1, consistent with the predominant role of adipose tissue-related dyslipidemia in this disease^8^, whereas cardiac arrhythmia and diseases of the veins were only increased by PC1. BMI is also a known risk factor for prolapsed disc^30^, though it is unknown whether this occurs due to mechanical overstraining, dysregulation of the metabolic and immune systems, or a combination of these^31^. That PC1 alone increased the risk of prolapsed disc strongly supports the role of mechanical stress over any form of lipotoxicity.

The decreased SES associated with PC2 is consistent with what has been reported for BMI^32, 33^, although recent results suggest that this is due to residual population stratification and non-genetic familial effects^34, 35^. Many other differences in lifestyle can be considered as concurrent with this change in SES (e.g. reduced education and income) whereas others are likely secondary to these (e.g. alcohol taken outside of meals^36^ and increased consumption of cheaper meats). Despite a moderate association with lower SES, PC1 was not associated with any of these secondary/concurrent changes, though the increased time spent using a computer may be indicative of white collar occupations rather than the physical jobs associated with PC2. The similarity of the effects of both PCs on smoking and alcohol consumption suggests mechanisms independent of SES. For example, increased smoking may reflect the use of smoking as a strategy for weight loss, leveraging the appetite suppressant effects of nicotine^37^, and is consistent with the observational correlation found between the number of cigarettes smoked and the risk of obesity^38^. Although we cannot exclude residual population stratification, the increased alcohol consumption found for both PC1 and PC2 suggests a directionality of causal effects which is rarely considered, as obesity is typically viewed as a consequence of alcohol consumption rather than the cause^39^. The lack of robust genetic instruments to explore the distinct effect of different alcoholic beverages render MR analysis suboptimal to resolve such reverse causations. The increased variation in diet and salt added to food may reflect a greater desire for palatable foods, possibly due to altered or reduced activation of the reward system which has been shown to occur in obese individuals (reviewed in 40). The healthy aspects of the dietary shift from PC1, e.g. greater consumption of fruits and vegetables, could possibly represent attempts to eat healthier, however this is likely to be highly confounded by biased reporting^41^. The weak to non-existent effects of cholesterol and other health risks or diseases on these measures of body size suggests that any lifestyle changes upon disease diagnosis are generally minimal and likely insufficient to have a real impact.

PC3, i.e. predisposition to abdominal fat deposition, was of particular interest as a hypothesis-free emergent component affecting body shape while remaining independent of its size or composition. This is in contrast to WHR which is highly correlated with BMI and body fat percentage, particularly in men^42^. The increased waist circumference at the expense of that of the hips suggests a change in body fat distribution, with a shift from subcutaneous (contributing to both hips and waist) to visceral adipose tissue (contributing to waist alone). This is supported by the enrichment for terms related to energy homeostasis and adipose tissue function, similar to those previously reported for WHR adjusted for BMI^43^, and the impacts on health which appear specific to lipotoxicity and are similar to what has been found for genetic variants affecting subcutaneous-to-visceral adipose tissue ratio^12, 13^. The implication of genes expressed in adipose tissues, particularly subcutaneous, is in line with the hypothesis that adipose tissue function, in particular its expandability, may be a critical factor in determining the distribution and impact of excess adiposity^7^ and its study may provide pharmaceutical avenues which may reduce health risks associated with obesity. The enrichment of both PCs 3 and 4 (as well as WHR) for genes expressed in several tissues of the digestive tract is not entirely surprising^44^, however their strong enrichment for all female-specific tissues tested was unexpected. It is possible that these tissues, and the genes expressed therein or associated sexual hormones, contribute to this phenotype, which could explain the increased variance of PC3 (and more generally WHR) observed in women compared to men. Additional studies will be required to determine how these genes and tissues are involved in the regulation of body fat distribution.

The additivity implied by the independence of the PCs’ causal effects provides a straightforward estimation of disease risk through linear combination, which proved more accurate than single trait-based prediction. Although causal effects are not ideally suited for within-sample outcome prediction, since they do not take advantage of non-causal correlates, their basis in causality proved reliable even out-of-population. The reason for the drop in accuracy seen for out-of-population PC- and BMI-based prediction of diabetes is unclear, though a number of factors may contribute, such as ethnicity-dependent differences in susceptibility, lifestyle, or ascertainment bias^45^, which exceed the scope of this study.

The systematic application of MR over a broad range of phenotypes carries the risk of violating its assumptions. Despite the steps taken to avoid any such violations or mitigate their impact, some sources of bias may remain inherent to the methods employed. GWAS effect estimates themselves have been shown to be biased due to population effects, such as parental effects, population stratification, and assortative mating^46^. Although such biases may be present in the exposure, they would only bias MR estimates if they affect the outcome in a similar fashion^34, 35^, which is difficult to test in a real setting. This study is also subject to other limitations of the MR approach, such as the estimated univariable causal effects summarizing the consequences of a lifetime exposure of a difference in phenotype at the population level. For example, the effect of 1 SD difference in weight (∼14kg) in the population is not equivalent to a sole gain/loss of 14kg, since this difference in the population distribution will be accompanied by a difference in average height as well (as illustrated by PC1 loadings). The causal effect being modeled linearly, it also averages what may be different effects in different strata of the population, i.e. if the causal effect is non-linear. For example, the effects of BMI on all cause mortality have been shown to be non-linear^47^, although the linearity of effects on specific diseases such as diabetes remains unclear^48^. In the case of non-linear effects, the MR causal effect estimates can be interpreted as an average effect at the population mean.

In summary, we established a scale-preserving method for the linear combination of traits at the summary-statistics level, mimicking a GWAS performed on a standardized composite trait but circumventing the need for individual-level data. Furthermore, this allows the combination of traits across different cohorts, since the covariance of effect size estimates between different traits can be estimated from summary statistics via cross-trait LD score regression intercept^49^. We also showed that cross-sex MR can reduce the bias by avoiding sample overlap, while preserving much of the statistical power from a potential one-sample MR using the full sample, still relying only on publicly available summary statistics. Applying these to the principal components of anthropometric traits, body size, adiposity, abdominal fat distribution, and lean mass, we showed that their distinct effects on health- and lifestyle-related outcomes can aid in understanding the etiology of the consequences of obesity. These components can be visualized and their effects on hundreds of outcomes compared through the shiny app which can be downloaded following the instructions at http://wp.unil.ch/sgg/pca-mr/. Finally, we showed their effectiveness in predicting obesity-related diseases, opening new avenues to classify individuals into more fine-scale obesity subtypes and identify personalized health risks. Although we identified four principal components here, larger sample sizes or more detailed anthropometric traits may achieve a finer scale identification of obesity subtypes.

## Methods

### Data and phenotype selection

We used GWAS summary statistics data derived from the UK Biobank^16^, a cohort of approximately 500,000 participants aged 37–73 (median 58), recruited between 2006 and 2010. Phenome-wide GWAS in UK Biobank data were performed and the summary statistics made available by the Neale Lab (http://www.nealelab.is/uk-biobank) for 3 different sex groups: men, women, and both sexes combined. This includes effect estimates for 13.7 million SNPs which were tested for association with 4,203 unique phenotypes across 361,194 unrelated, white British individuals (167,020 men, 194,174 women). All were adjusted for age, age^2^, sex, age*sex, age^2^*sex, and the top 20 genetic PCs to correct for population stratification. We excluded SNPs from the HLA region (chr6: 28,477,797–33,448,354, www.ncbi.nlm.nih.gov/grc/human/regions/MHC?asm=GRCh37). For continuous phenotypes, we used effect estimates for the inverse rank-normalized trait. Summary statistics for WHR were calculated in the UK Biobank across a similar sample of 378,139 unrelated, white British individuals (175,155 men, 202,984 women) and correcting for the same covariates after inverse rank-normalization. For binary traits, we divided the effect estimates and standard errors by the square root of the variance of the trait (that of the analyzed sample where provided, otherwise across a similar subset of the UK Biobank). As such, all effects are expressed on the standard deviation (SD) scale and comparable with continuous traits. The linear models used to estimate the SNP-binary trait association provide well calibrated p-values, as long as rare SNPs (MAF < 0.1%) are not evaluated for a trait with highly imbalanced case fraction (< 10%) or as long as the product of MAF and disease prevalence exceeds 0.0001 ^50^. Since we analyzed diseases only with >1% prevalence, any SNPs with MAF>1% are safe to use as instruments. he largest fraction of low-frequency (MAF<1%) IVs were observed for height (88/1704 SNPs, i.e. 5.2% vs. max 2.8% for other traits/PCs) and the lowest disease prevalence were between 1–2%. Still the resulting causal effects from MR analysis between height and these diseases did not change appreciably upon the exclusion of IVs below 1% MAF (Supplementary Figure 122).

In total, 232 phenotypes that had at least one GWS SNP (p < 5*10^-8^) were divided into 5 mutually exclusive categories: body measures (14 phenotypes), continuous measures of health (37 phenotypes), dietary habits (19 phenotypes), diseases (108 phenotypes), and lifestyle factors (54 phenotypes), which are briefly described below. The full list of selected phenotypes can be found in **Supplementary table 1**.

Body measures included BMI, height, weight, hip and waist circumference and WHR, as well as bioimpedance-derived fat and lean mass estimates in arms, legs, trunk, and overall body fat percentage. For arms and legs, summary statistics were available for left and right sides. As these were almost identical, the statistics for the left side alone were used. Basal metabolic rate was also included in body measurements as it is derived from bioimpedance measures.

Continuous health outcomes included the biomarker panel of the UK Biobank, including 34 biomarkers measured in either blood or urine, as well as systolic and diastolic blood pressure (BP), heart rate at rest and during effort, and forced vital capacity (FVC).

Dietary habits were obtained from a food frequency questionnaire (FFQ). UK Biobank also includes more specific questions such as 24-hour recall (food consumed in the last day) which is more reliable but less representative, as well as specific questions such as the type of bread or milk typically consumed, however these were not included in the present analysis.

Disease summary statistics in the UK Biobank were available for self-reported disease status, ICD-10-classified hospital diagnoses, and diseases curated by the Neale Lab in collaboration with the FinnGen consortium (www.finngen.fi). We included data from both self-reported answers and the FinnGen curated diseases, excluding the raw ICD10 diagnoses which were considered less informative. We included any diseases which had a prevalence of at least 1% in the analyzed sample.

Lifestyle included both environmental factors and lifestyle choices, mainly relating to physical activity, alcohol consumption, smoking, sleep, work, and socio-economic status.

We restricted SNPs to those in common between the summary statistics from Neale and the UK10K reference panel used for LD pruning. We also removed SNPs with a minor allele frequency below 0.001, resulting in 9,675,947 SNPs. For each trait, SNPs were pruned separately using *plink* v1.90b6 with the UK10K European LD panel to obtain independent MR instruments. SNPs were considered independent if separated by more than 10Mb or the linkage disequilibrium was r^2^ < 0.01.

### High-accuracy body composition measurements

In addition to bioimpedance measurements, the UK Biobank provides other body composition phenotypes derived from more accurate methods, namely DXA and MRI. Unfortunately, the sample sizes for these phenotypes were too low (∼5,000 individuals) for their inclusion as body phenotypes in the PCA. We did, however, investigate whether approximating these traits through linear combination of other available traits (with regression weights calculated in the UK Biobank) could provide a hypothesis-driven alternative to the hypothesis-free PCA approach. We were able to estimate 57 body composition measurements using the same 14 anthropometric and bioimpedance-based traits with varying accuracy (see Supplementary Table 2). 18 out of 57 had an r^2^ above 80%, including abdominal SAT, and another 18 had r^2^ above 70%, including VAT. Others, such as bone mass and liver fat percentage could not be accurately approximated using the included traits.

### Principal component analysis

Principal component analysis (PCA) was performed on a matrix of effect estimates of independent SNPs on the 14 body traits described above. We selected all SNPs with a genome-wide significant (GWS) effect (p < 5*10^-8^) on at least one of the body traits and pruned them using the same procedure as for single traits (distance > 10Mb or r^2^ < 0.01). SNPs were prioritized according to the highest rank within any significantly associated trait, i.e. for each trait, all SNPs significantly associated with it were ranked by p-value; then each SNP was attributed a priority based on the highest rank obtained with any significantly associated trait. This was done to avoid the well-powered traits (e.g. height) overshadowing other traits and driving the SNP selection. Any missing effect estimates in the resulting matrix were set to 0. Data was neither centered nor scaled prior to the PCA, as the effect estimates were standardized and should therefore have zero mean and their variance for a given trait is informative (distantly related to trait heritability).

PCA was then performed on the resulting matrix of genetic effects to obtain the PC loadings, which were then used to calculate SNP-PC associations across the entire genome. These were then individually pruned to obtain the final set of SNPs for each PC.

Although the SNP-phenotype associations are standardized (effects are on an SD/SD scale), a linear combination of effects would yield non-standard effects since the resulting traits do not have a variance of 1. To maintain comparable effects between PCs and traits, the PC loadings should be scaled such that the resulting βs would represent SNP effects on composite traits with unit variance, which would typically involve performing the rotation on the phenotypes, standardizing, and rerunning a GWAS on the resulting composite trait with variance 1. Instead, we can consider the linear regression coefficients for trait *j* calculated as:

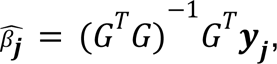

where *y_j_* is the vector of values for trait *j* and *G* is the genotype matrix, and a weighted sum of *n* variables:

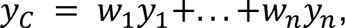

where *w_j_* is the weight for trait *j.* The effect size for the composite trait can then be calculated as:

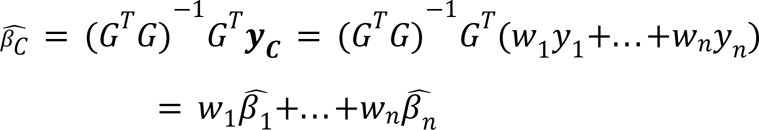

The expected variance of variable *y_C_* can be written as^51^

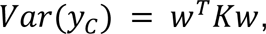

where *K* is the covariance matrix of the *n* traits composing *y_C_*, which in the case of standardized trait variables, is the correlation matrix. Knowing the variance of the composite trait, we can simply rescale the vector of weights such that the expected variance in the composite output variable *y_C_* is equal to one, resulting in standardized effect sizes. Applying this to the PC loadings, we rescaled the weights as follows:

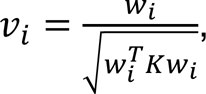

where *w_i_* and *v_i_* are the unadjusted and adjusted (trait) loading vectors for PC i, respectively, and K is the pairwise phenotypic correlation matrix of body trait phenotypes, which was calculated on a subset of the UK Biobank similar to that used for the summary statistics (i.e. unrelated white British individuals) and the phenotypes were corrected for the same covariates as used by the Neale lab prior to calculating the correlation. The effect estimates were then calculated using these adjusted loadings, i.e.

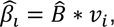

where 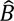 is the matrix of genetic effects on the 14 anthropometric traits. The corresponding standard errors were calculated following the formula for a weighted sum of random variables^51^:

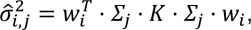

where 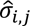 is the standard error for the association of SNP j with PC i and Σ_j_ is a diagonal matrix with the standard error of the association of SNP j with each trait. Since we use only UK Biobank summary statistics, the correlation between effect estimates simplifies to the phenotypic correlation between the traits. The advantage of this approach is that we do not need to calculate the composite trait and run a GWAS but can directly compute the association summary statistics.

### Tissue specificity and pathway enrichment

We tested PC- and trait-associated SNPs for enrichment of tissue-specific genes using the MAGMA tool for gene set analysis^17^ through the FUMA v 1.3.5e interface^18^ using the default parameters, with a Bonferroni-adjusted p-value threshold. We used gene expression data in 54 tissues from GTEx v8^19^ as well as brain development and age data from BrainSpan^20^.

We used the molecular pathway gene sets defined by DEPICT^21^ and tested for enrichment using PASCAL^22^. As with all pathway enrichment methods, we assume that most regulatory variants for a given gene are in close (50kb) physical proximity to the gene body, hence we may ignore more distant regulatory variants.

### Mendelian randomization

We used inverse-variance weighted (IVW) Mendelian randomization (MR) to estimate causal effects of body traits and PCs on all non-body traits, as well as the reverse. MR mimics a randomized controlled trial (RCT) where the treatment corresponds to the random allocation of an exposure-associated allele, called an instrumental variable (IV)^52^. MR relies on three key assumptions to infer causality: 1, the IV is associated with a change in the exposure; 2, the IV is independent of the outcome, except through its association with the exposure; 3, the IV is independent of any confounders of the exposure-outcome association. If these assumptions are verified, MR provides an unbiased estimate of the causal effect of the exposure on the outcome and can be done using summary statistics data alone. We used IVW MR as default method for all our analyses, but we compared IVW causal effect estimates to those obtained from weighted median-based MR to ensure robustness.

### Cross-sex Mendelian randomization

In our case, the use of summary data from the UK Biobank for both the exposure and outcome effect sizes (i.e. full sample overlap) would lead to a bias in MR causal effect estimate in the direction of the observed correlation of the phenotypes^53^. To circumvent this, we used the existing summary statistics for two non-overlapping samples from the Neale Lab, those for men and women separately. Each sex was used as exposure on the other as outcome and then both causal effect estimates were meta-analyzed (using inverse variance weighting). This removed the correlation between the error terms of the effect estimates of IVs on exposure and outcome, significantly reducing the bias from sample overlap, while minimizing loss of power. This would lead to a slight bias away from zero even when there is no true difference in the genetic effects on the exposure in men and women, given that:

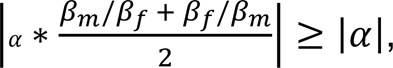

where ⍺ is the common causal effect (for men and women), β_m_ and β_f_ are the coefficients of association for a given IV with the exposure for men and women, respectively. This bias increases if there is a difference between sexes in the strength of association with the exposure, but its magnitude remains small. For example, if the SNP effects on the exposure were consistently 1.37 times stronger in one sex, the relative theoretical bias would only be 5%. In practice, however, the IVs were filtered for genome-wide significance in each sex prior to being used as exposure, resulting in fewer IVs being selected in the sex with weaker effects (on the exposure). This reduced power and increased variance in the overestimated causal effect, which was correspondingly down-weighted by the IVW meta-analysis. For example, we found PC3 to have a strong sex-specificity, with effects on exposure tending to be ∼2.2 times stronger in women, which would theoretically lead to a 34% bias away from the null. However, this yielded 242 GWS IVs in women but only 33 in men. This increased the variance in the male exposure - female outcome causal effect estimate by a median of 6.8-fold, resulting in a corresponding down-weighting of the overestimated causal effect. In this example, we expect a meta-analyzed estimate which is biased towards the null by ∼32% (since the IVW causal effect estimate is expected to be (((6.1/7.1)*(1/1.7) + (1/7.1)*1.7)\**α*=0.68\**α*). Importantly, since the bias is multiplicative, this introduces no bias in the absence of causal effect, indicating it will not affect the type I error rate.

Although the PCs were constructed using combined-sex summary statistics, the selected IVs were filtered for genome-wide significance in the sex-specific summary statistics prior to MR analysis to avoid weak instrument bias inflating causal effect estimates. This may slightly exacerbate Winner’s curse, inflating the SNP-exposure association, which would result in a small bias towards the null. In the presence of a true causal effect, however, the SNP-outcome association (being assessed in the same sample) may be proportionally increased, which would mitigate this bias. Such biases are difficult to correct for without using 3 independent samples.

For each sex, the GWS IVs associated with the exposure formed the initial set of IVs. Those with significantly larger effects on the outcome than on the exposure were removed, as these would indicate a violation of MR assumptions (likely reverse causality and/or confounding). The effect sizes being on a standardized scale, they were compared directly using a one-sided t-test and removed if the magnitude of the effects in the outcome were significantly greater (p < 0.05). To avoid unreliable causal effect estimates, MR was only performed if at least 10 IVs remained. This was done using the *TwoSampleMR* R package v0.5.4^54^.

The MR causal effect estimates from individual IVs were tested for heterogeneity using Cochran’s Q test:

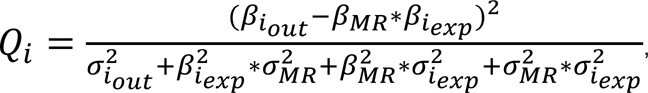

where β_iexp_ and β_iout_ are the coefficients of association of SNP i with the exposure and outcome, respectively, with σ_iexp_ and σ_iout_ the corresponding standard errors, and β_MR_ and σ_MR_ are the MR estimate and standard error of the causal effect of the exposure on the outcome. The test statistic Q_i_ follows a *χ*^2^ distribution with 1 degree of freedom. If any of the IVs used had an associated p < 10^-3^, the most heterogeneous one (with the lowest p-value) was removed. If at least 5 IVs remained, the MR was then repeated with the remaining IVs. In practice, few SNPs were filtered as outliers (on average less than 1%), though we also explored alternative methods for dealing with heterogeneity, namely performing no filtering, using weighted median MR with or without filtering, or using exact Q statistics^55^ instead of Cochran’s Q (see Supplementary figures 123–128). All of these produced similar effect estimates, though those from weighted median MR tended to be ∼6% lower and lacked the statistical power to find many effects.

Due to the standardized scales of IV effect estimates, the resulting causal effects are on a scale representing the SD change in the outcome for a change of 1 SD in the exposure.

To account for multiple testing, we adjusted the p-value threshold with Bonferroni correction for the number of tests within the exposure-outcome category pair. For example, the p-value threshold for the effect of BMI (body phenotype) on T2D (disease) is 0.05 / (number of body phenotypes * number of diseases).

### Sex-specific Mendelian randomization

In most cases, we were instead able to obtain unbiased causal effect estimates using the exposure summary statistics from the opposite sex on those of the sex of interest in the outcome. This relies on the strength of the association between the IVs and the exposure to be identical (or at least not systematically different) between sexes. We tested this using both paired Wilcoxon signed-rank test on the absolute genetic effects of the IVs and total least squares (TLS) regression. Exposures with non-significant results for both tests were considered identical in both sexes for this purpose and the summary statistics for the opposite sex were used for the exposure.

### Comparison of causal effects

To test the significance of a difference in effects of two exposures on a given outcome, we used a two-sided Z-test^56^. We accounted for multiple testing using Bonferroni correction, adjusting for the total number of tests, i.e. the number of significant effects of either exposure on the category outcomes. For example, to test whether the effects of BMI on T2D were different from those of WHR, the significance threshold would be 0.05 / total number of diseases significantly affected by either BMI or WHR.

In some cases, it was useful to compare the causal effects of two exposures on all outcomes of a category (e.g. the effects of weight on disease risk compared to those of PC1) or the same exposure in different experimental settings (comparison of methods or sex-stratified analyses). In these cases, we obtained the slope estimate using TLS regression with no intercept, which considers error in both axes rather than ordinary least squares (OLS) which minimizes only the vertical offset. The standard error on the angle of the regression line (rather than the slope, which is not symmetrical and dependent on the phenotype placed on the y-axis) was computed using a jackknife procedure, which was then used to compare the obtained estimate with the null hypothesis that the true causal effects were identical (i.e. an angle of 45° or a slope of 1). The TLS procedure was performed using the *deming* R package v1.4^57^.

### Prediction of disease risk

The accuracy of disease prediction was assessed in-sample in the UK Biobank across 371’523 unrelated, white British individuals (199,699 women, 171,824 men) for diabetes, high cholesterol, and hypertension. PCs were calculated based on the 14 scaled and centered anthropometric traits for each individual. These were then combined in a disease-specific linear combination based on the estimated causal effect on the disease. Rather than ignoring PCs with non-significant effects, the effect estimates were inverse variance weighted to account for the uncertainty of the effect:

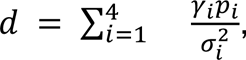

where **γ*_i_* and *σ_i_* are the estimated causal effect of PC i on the disease of interest and its standard error, respectively, and *p_i_* is the individual’s phenotypic realization of that PC.

The accuracy of this predictor was assessed using receiver operating characteristic (ROC) curves by comparing the area under the curve (AUC) with those of BMI and WHR. The significance of the difference between the AUCs for each predictor was determined using DeLong’s test in the *pROC* R package (version 1.16.2)^58^.

Out-of-sample prediction accuracy was assessed in a sample of 76,756 UK Biobank participants (42,407 women, 34,349 men) who were not flagged as “in.white.British.ancestry.subset” in the sample QC file. This includes individuals who either did not self-report ‘white British’ or whose genetic ancestry (as determined by the genomic PCs) was dissimilar from other white British individuals. This sample thereby excluded all the individuals included in the original analysis and any relatives in the UK Biobank. Simulating a clinical setting, the phenotypes were scaled according to the original distribution, i.e. the mean and SD of the original sample were used to scale individuals without relying on the distribution in the second sample. Using the values of the second distribution for scaling changed little (the AUC increased by at most 4.4*10^-4^).

## Data Availability

Data was provided by the UK Biobank and the Neale lab.

https://wp.unil.ch/sgg/pca-mr/

## Acknowledgements

This research has been conducted using the UK Biobank resource (#16389), which has been approved by the National Research Ethics Service Committee. The computations have been carried out on the HPC server of the Lausanne University Hospital. Z.K. was funded by the Swiss National Science Foundation (31003A-143914 and 310030-189147). B.D. is supported by the Swiss National Science Foundation (NCCR Synapsy, project grant Nr. 32003B_135679, 32003B_159780, 324730_192755 and CRSK-3_190185) and the Leenaards Foundation. LREN is very grateful to the Roger De Spoelberch and Partridge Foundations for their generous financial support. T.O.K. was funded by the Novo Nordisk Foundation (NNF17OC0026848, NNF18CC0034900).

## Author contributions

A.S. and J.S. preprocessed the data and performed the initial analyses. N.M. assisted with initial data preprocessing. B.D. contributed to the initial design. J.S. performed the remaining analyses. Z.K. and J.S. developed the methods. Z.K. designed and supervised the project. J.S. wrote the initial manuscript. All authors provided critical feedback, helped interpret the findings and revise the manuscript.

## References

1. Boles, A., Kandimalla, R. & Reddy, P. H. Dynamics of diabetes and obesity: Epidemiological perspective. Biochim. Biophys. Acta Mol. Basis Dis. 1863, 1026–1036 (2017).

2. Ortega, F. B., Lavie, C. J. & Blair, S. N. Obesity and Cardiovascular Disease. Circ. Res. 118, 1752–1770 (2016).

3. Pischon, T. et al. General and abdominal adiposity and risk of death in Europe. N. Engl. J. Med. 359, 2105–2120 (2008).

4. Kaess, B. M. et al. The ratio of visceral to subcutaneous fat, a metric of body fat distribution, is a unique correlate of cardiometabolic risk. Diabetologia 55, 2622–2630 (2012).

5. Rosenquist, K. J. et al. Visceral and subcutaneous fat quality and cardiometabolic risk. JACC Cardiovasc. Imaging 6, 762–771 (2013).

6. Abraham, T. M., Pedley, A., Massaro, J. M., Hoffmann, U. & Fox, C. S. Association between visceral and subcutaneous adipose depots and incident cardiovascular disease risk factors. Circulation 132, 1639–1647 (2015).

7. Virtue, S. & Vidal-Puig, A. Adipose tissue expandability, lipotoxicity and the Metabolic Syndrome--an allostatic perspective. Biochim. Biophys. Acta 1801, 338–349 (2010).

8. Heymsfield, S. B. & Wadden, T. A. Mechanisms, Pathophysiology, and Management of Obesity. The New England journal of medicine vol. 376 1492 (2017).

9. Zhang, M., Hu, T., Zhang, S. & Zhou, L. Associations of Different Adipose Tissue Depots with Insulin Resistance: A Systematic Review and Meta-analysis of Observational Studies. Sci. Rep. 5, 18495 (2015).

10. Porter, S. A. et al. Abdominal subcutaneous adipose tissue: a protective fat depot? Diabetes Care 32, 1068–1075 (2009).

11. Ried, J. S. et al. A principal component meta-analysis on multiple anthropometric traits identifies novel loci for body shape. Nat. Commun. 7, 13357 (2016).

12. Yaghootkar, H. et al. Genetic evidence for a normal-weight ‘metabolically obese’ phenotype linking insulin resistance, hypertension, coronary artery disease, and type 2 diabetes. Diabetes 63, 4369–4377 (2014).

13. Yaghootkar, H. et al. Genetic Evidence for a Link Between Favorable Adiposity and Lower Risk of Type 2 Diabetes, Hypertension, and Heart Disease. Diabetes 65, 2448–2460 (2016).

14. Lotta, L. A. et al. Integrative genomic analysis implicates limited peripheral adipose storage capacity in the pathogenesis of human insulin resistance. Nat. Genet. 49, 17–26 (2017).

15. Ji, Y. et al. Genome-Wide and Abdominal MRI Data Provide Evidence That a Genetically Determined Favorable Adiposity Phenotype Is Characterized by Lower Ectopic Liver Fat and Lower Risk of Type 2 Diabetes, Heart Disease, and Hypertension. Diabetes 68, 207– 219 (2019).

16. Sudlow, C. et al. UK biobank: an open access resource for identifying the causes of a wide range of complex diseases of middle and old age. PLoS Med. 12, e1001779 (2015).

17. de Leeuw, C. A., Mooij, J. M., Heskes, T. & Posthuma, D. MAGMA: generalized gene-set analysis of GWAS data. PLoS Comput. Biol. 11, e1004219 (2015).

18. Watanabe, K., Taskesen, E., van Bochoven, A. & Posthuma, D. Functional mapping and annotation of genetic associations with FUMA. Nat. Commun. 8, 1826 (2017).

19. Aguet, F. et al. The GTEx Consortium atlas of genetic regulatory effects across human tissues. bioRxiv 787903 (2019) doi:10.1101/787903.

20. Miller, J. A. et al. Transcriptional landscape of the prenatal human brain. Nature 508, 199– 206 (2014).

21. Pers, T. H. et al. Biological interpretation of genome-wide association studies using predicted gene functions. Nature Communications vol. 6 (2015).

22. Lamparter, D., Marbach, D., Rueedi, R., Kutalik, Z. & Bergmann, S. Fast and Rigorous Computation of Gene and Pathway Scores from SNP-Based Summary Statistics. PLOS Computational Biology vol. 12 e1004714 (2016).

23. Lang, J. et al. Association of serum albumin levels with kidney function decline and incident chronic kidney disease in elders. Nephrol. Dial. Transplant 33, 986–992 (2018).

24. Marra, M. et al. Assessment of Body Composition in Health and Disease Using Bioelectrical Impedance Analysis (BIA) and Dual Energy X-Ray Absorptiometry (DXA): A Critical Overview. Contrast Media Mol. Imaging 2019, 3548284 (2019).

25. Locke, A. E. et al. Genetic studies of body mass index yield new insights for obesity biology. Nature 518, 197–206 (2015).

26. van der Klaauw, A. A. & Farooqi, I. S. The hunger genes: pathways to obesity. Cell 161, 119–132 (2015).

27. Halban, P. A. et al. β-cell failure in type 2 diabetes: postulated mechanisms and prospects for prevention and treatment. Diabetes Care 37, 1751–1758 (2014).

28. Duclos, M. Osteoarthritis, obesity and type 2 diabetes: The weight of waist circumference. Ann. Phys. Rehabil. Med. 59, 157–160 (2016).

29. Hyppönen, E., Mulugeta, A., Zhou, A. & Santhanakrishnan, V. K. A data-driven approach for studying the role of body mass in multiple diseases: a phenome-wide registry-based case-control study in the UK Biobank. Lancet Digit Health 1, e116–e126 (2019).

30. Schroeder, G. D., Guyre, C. A. & Vaccaro, A. R. The epidemiology and pathophysiology of lumbar disc herniations. Semin. Spine Surg. 28, 2–7 (2016).

31. Weiler, C. et al. Histological analysis of surgical lumbar intervertebral disc tissue provides evidence for an association between disc degeneration and increased body mass index. BMC Res. Notes 4, 497 (2011).

32. Wang, Y. & Beydoun, M. A. The Obesity Epidemic in the United States Gender, Age, Socioeconomic, Racial/Ethnic, and Geographic Characteristics: A Systematic Review and Meta-Regression Analysis. Epidemiologic Reviews vol. 29 6–28 (2007).

33. Howe, L. D. et al. Effects of body mass index on relationship status, social contact and socio-economic position: Mendelian randomization and within-sibling study in UK Biobank. Int. J. Epidemiol. (2019) doi:10.1093/ije/dyz240.

34. Darrous, L., Mounier, N. & Kutalik, Z. Simultaneous estimation of bi-directional causal effects and heritable confounding from GWAS summary statistics. Genetic and Genomic Medicine (2020) doi:10.1101/2020.01.27.20018929.

35. Brumpton, B. et al. Avoiding dynastic, assortative mating, and population stratification biases in Mendelian randomization through within-family analyses. Nat. Commun. 11, 3519 (2020).

36. Oksanen, A. & Kokkonen, H. Consumption of Wine with Meals and Subjective Well-being: A Finnish Population-Based Study. Alcohol Alcohol 51, 716–722 (2016).

37. Mineur, Y. S. et al. Nicotine decreases food intake through activation of POMC neurons. Science 332, 1330–1332 (2011).

38. Dare, S., Mackay, D. F. & Pell, J. P. Relationship between smoking and obesity: a cross-sectional study of 499,504 middle-aged adults in the UK general population. PLoS One 10, e0123579 (2015).

39. Traversy, G. & Chaput, J.-P. Alcohol Consumption and Obesity: An Update. Curr. Obes. Rep. 4, 122–130 (2015).

40. Stice, E., Spoor, S., Ng, J. & Zald, D. H. Relation of obesity to consummatory and anticipatory food reward. Physiol. Behav. 97, 551–560 (2009).

41. Pirastu, N. et al. Using genetics to disentangle the complex relationship between food choices and health status. bioRxiv 829952 (2019) doi:10.1101/829952.

42. Sulc, J., Winkler, T. W., Heid, I. M. & Kutalik, Z. Heterogeneity in Obesity: Genetic Basis and Metabolic Consequences. Curr. Diab. Rep. 20, 1 (2020).

43. Shungin, D. et al. New genetic loci link adipose and insulin biology to body fat distribution. Nature 518, 187–196 (2015).

44. Winkler, T. W. et al. A joint view on genetic variants for adiposity differentiates subtypes with distinct metabolic implications. Nat. Commun. 9, 1946 (2018).

45. Goff, L. M. Ethnicity and Type 2 diabetes in the UK. Diabet. Med. 36, 927–938 (2019).

46. Young, A. I. et al. Mendelian imputation of parental genotypes for genome-wide estimation of direct and indirect genetic effects. bioRxiv 2020.07.02.185199 (2020) doi:10.1101/2020.07.02.185199.

47. Sun, Y.-Q. et al. Body mass index and all cause mortality in HUNT and UK Biobank studies: linear and non-linear mendelian randomisation analyses. BMJ 364, l1042 (2019).

48. Wainberg, M. et al. Homogeneity in the association of body mass index with type 2 diabetes across the UK Biobank: A Mendelian randomization study. PLoS Med. 16, e1002982 (2019).

49. Bulik-Sullivan, B. et al. An atlas of genetic correlations across human diseases and traits. Nat. Genet. 47, 1236–1241 (2015).

50. Loh, P.-R., Kichaev, G., Gazal, S., Schoech, A. P. & Price, A. L. Mixed-model association for biobank-scale datasets. Nat. Genet. 50, 906–908 (2018).

51. Johnson, A. R. & Wichern, D. W. Applied multivariate statistical analysis. Biometrics 44, 920 (1988).

52. Burgess, S., Butterworth, A. & Thompson, S. G. Mendelian randomization analysis with multiple genetic variants using summarized data. Genet. Epidemiol. 37, 658–665 (2013).

53. Burgess, S., Davies, N. M. & Thompson, S. G. Bias due to participant overlap in two-sample Mendelian randomization. Genet. Epidemiol. 40, 597–608 (2016).

54. Hemani, G. et al. The MR-Base platform supports systematic causal inference across the human phenome. eLife vol. 7 (2018).

55. Bowden, J. et al. Improving the accuracy of two-sample summary-data Mendelian randomization: moving beyond the NOME assumption. Int. J. Epidemiol. 48, 728–742 (2019).

56. Paternoster, R., Brame, R., Mazerolle, P. & Piquero, A. USING THE CORRECT STATISTICAL TEST FOR THE EQUALITY OF REGRESSION COEFFICIENTS. Criminology 36, 859–866 (1998).

57. Therneau, T. deming: Deming, Theil-Sen, Passing-Bablock and Total Least Squares Regression, R package version 1.4. https://CRAN.R-project.org/package=deming (2018).

58. Robin, X. et al. pROC: an open-source package for R and S+ to analyze and compare ROC curves. BMC Bioinformatics 12, 77 (2011).

